# Pre-diagnostic circulating untargeted metabolomics and risk of overall and clinically significant prostate cancer: A systematic review meta-analysis

**DOI:** 10.1101/2025.02.27.25321444

**Authors:** Harriett Fuller, Orietta P. Agasaro, Jim M. Guevera, Burcu F. Darst

## Abstract

**Background:** Metabolomic dysregulation contributes to prostate cancer (PCa) pathogenesis, and studies suggest that circulating metabolites have strong potential to act as clinical biomarkers. However, evidence of associations between circulating metabolites with overall and clinically significant PCa risk has not been quantitively aggregated.

**Methods:** We performed a systematic review and meta-analysis of untargeted pre-diagnostic circulating metabolomic studies across four clinically distinct outcomes: overall, low- to intermediate-risk, high- to very high-risk, and lethal PCa, each compared to controls.

**Results:** Twelve studies were identified in the systematic review, and up to 408 metabolites were meta-analyzed across the four PCa outcomes. Three, eleven and nineteen metabolites were significantly associated with risk of overall, high- to very high-risk and lethal PCa, respectively. Metabolites associated with high- to very high-risk PCa were significantly enriched for lipids. Limited evidence of correlation between metabolite effects across outcomes was identified, highlighting potentially unique metabolite drivers of high-risk and lethal PCa. In follow-up analyses, 13 of the significant metabolites were found to be drug and/or dietary modifiable.

**Conclusions:** These findings suggest the strong potential for metabolites to inform risk of lethal PCa, which could inform risk-stratified screening strategies and facilitate the identification of targets for PCa prevention.

## Introduction

Globally, prostate cancer (PCa) is the fifth leading cause of cancer mortality and second most frequently diagnosed cancer in men^1,2^. Between 2013-2019, in the US, survival rates were 34% for distant PCa compared to 97% for any PCa^3^, highlighting the need to improve our knowledge of aggressive PCa risk factors and pathogenesis. One promising avenue of PCa biomarker identification is metabolomics. Due to the increased energy demands of cellular malignancy^4,5^, metabolic dysregulation is expected to contribute to PCa pathogenesis. As such, metabolomic investigations could elucidate biological mechanisms that drive tumor development and progression and ultimately improve screening strategies and the identification of lethal PCa. Untargeted metabolomic investigations, which aim to characterize all metabolites in a sample in an unbiased manner, offer the greatest potential for discovery^6–8^.

Although metabolomic epidemiology investigations of PCa risk have been conducted, replication of metabolites identified has been challenging due to limited sample sizes and between-study heterogeneity, particularly regarding metabolomic platforms, data processing, and analysis procedures. Accordingly, three previous systematic reviews reported limited reproducibility across circulating metabolomic studies of PCa, but highlighted the potential role of select amino acids and lipids in PCa risk assessment and distinguishing aggressive from non-aggressive PCa at diagnosis^9–11^. However, metabolomic evidence has not been quantitatively aggregated across studies through meta-analyses, limiting the interpretability of findings, ability to assess heterogeneity, and strength of conclusions. Moreover, previous systematic reviews have not compared metabolite findings across clinically distinct PCa outcomes; identifying pre-diagnostic metabolites that are specific to high-risk or lethal PCa could have notable clinical implications for screening and preventive practices.

We conducted a systematic review and meta-analysis of untargeted pre-diagnostic circulating metabolomic studies to quantitatively evaluate current evidence and robustly identify metabolites associated with risk of overall, low- to intermediate-risk, high- to very high-risk and lethal PCa and to compare risk profiles across these clinically distinct PCa outcomes. Furthermore, we conducted a comprehensive bias assessment to investigate potential causes of heterogeneity present in PCa molecular epidemiological studies.

## Results

### Study Identification

Upon conducting a systematic review using PubMed and Embase through October 17^th^, 2024, 1,282 publications were identified for abstract screening (**Figure 1**, **Methods**). After limiting to prospective investigations with ≥50 metabolites measured in pre-diagnostic blood, 12 publications were eligible for inclusion, all of which were nested case-control studies. A total of 15,382 predominantly (>90%) European descent participants, including 7,643 cases and 7,739 controls, were included across 7 cohorts (Alpha-Tocopherol, Beta-Carotene Cancer Prevention [ATBC]^12–15^, European Prospective Investigation into Cancer and Nutrition [EPIC]^16,17^, Supplémentation en Vitamines et Minéraux Antioxydants [SU.VI.MAX]^18,19^, Health Professionals Follow-up Study [HPFS]^20^, Physicians’ Health Study [PHS]^20^, Northern Sweden Health and Disease Study [NSHDS]^21^, and Cancer Prevention Study-II Nutrition Cohort [CPS-11]^22^; **Table 1**, **Supplementary Tables 1-2**). Ten studies quantified metabolite levels via MS and two used NMR, with one study using both. The average number of metabolites reported across MS studies was 478 (range 67-1,100) and across NMR studies was 208 (range 159-277).

**Figure 1:**
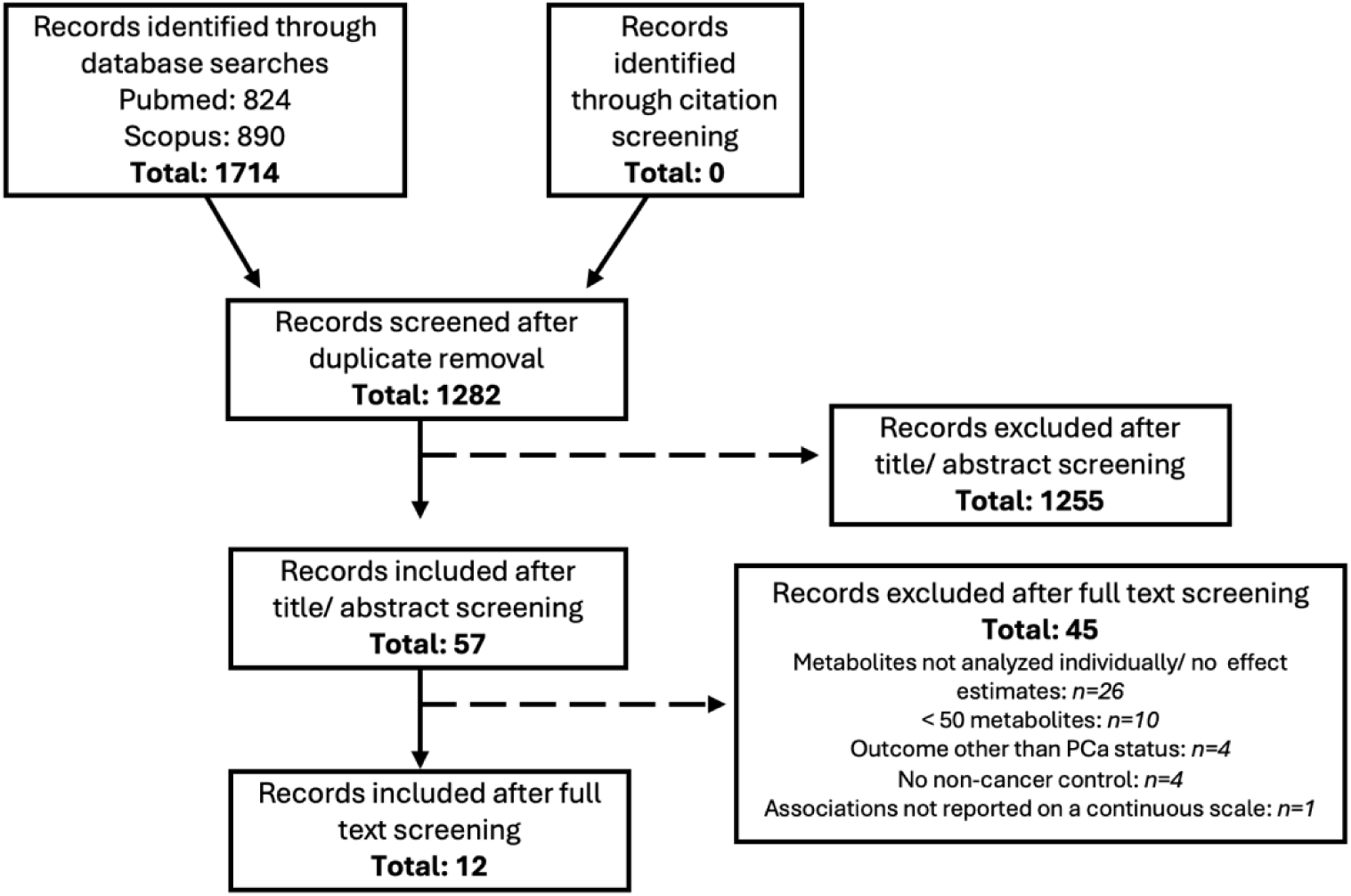
PRISMA flow diagram for systematic review of untargeted metabolomic studies and prostate cancer risk.

**Table 1:**
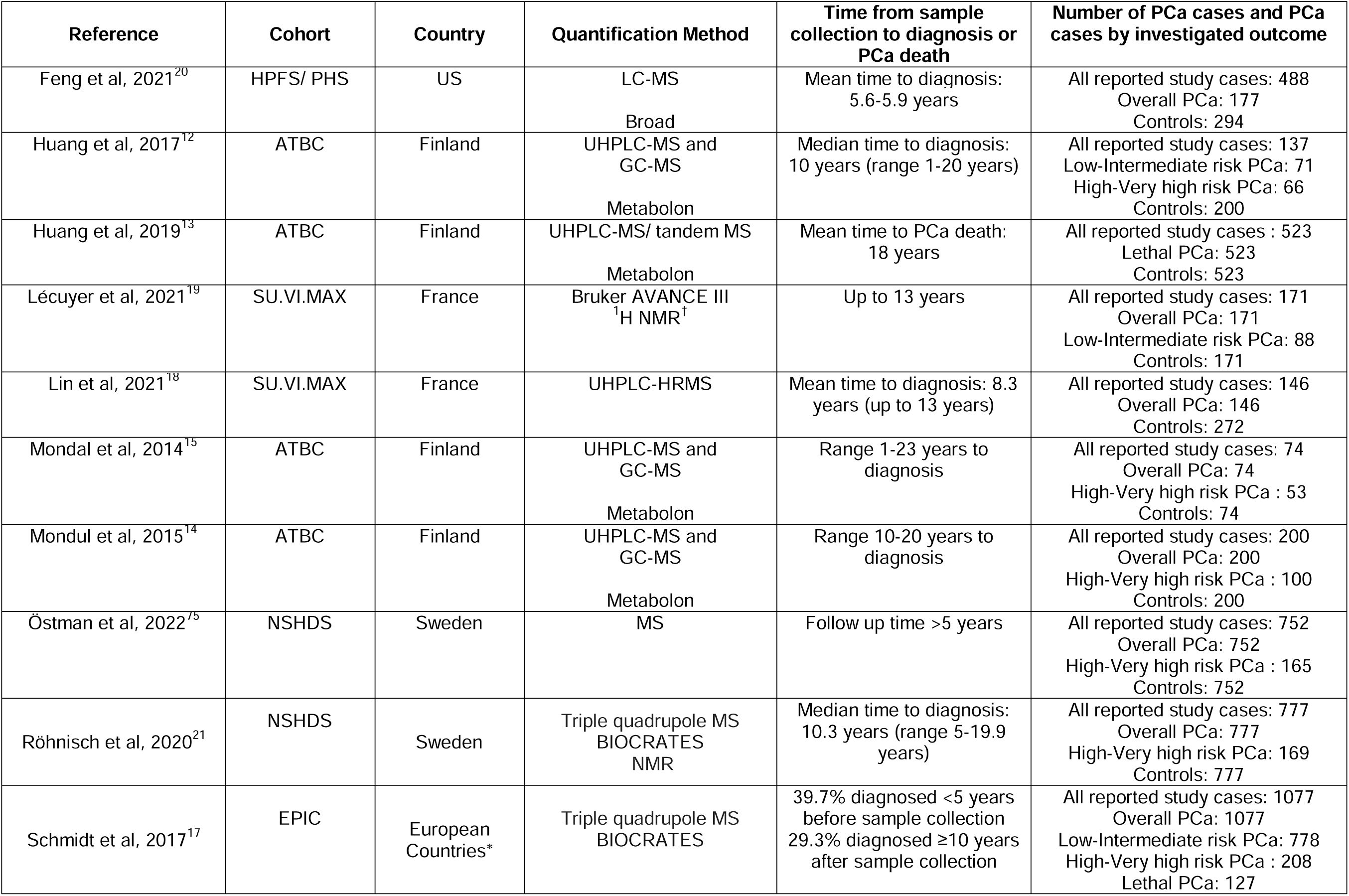
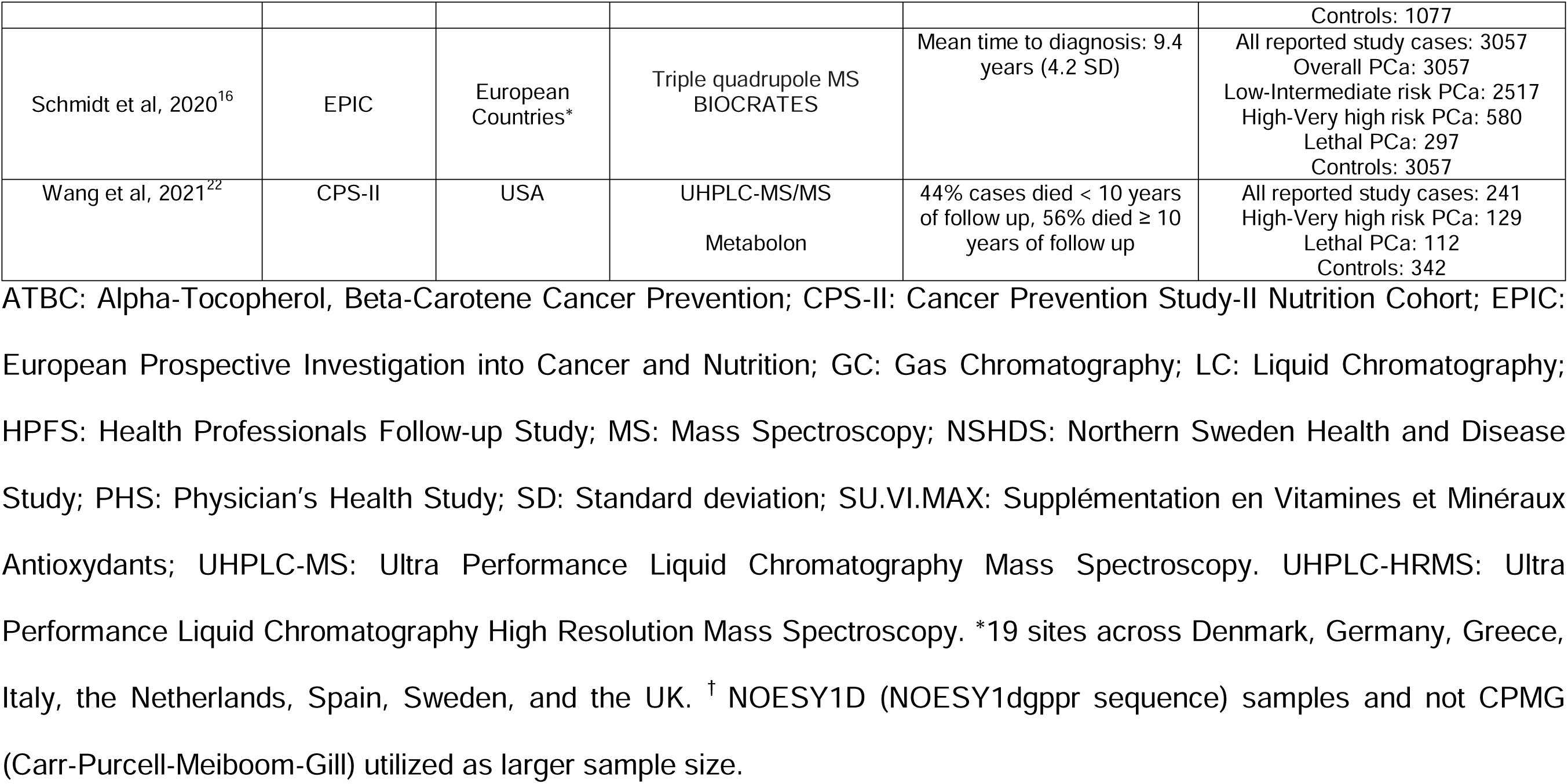
Description of studies included in the systematic review and meta-analysis.

We evaluated four PCa outcomes, informed by the National Comprehensive Cancer Network (NCCN)^23^: overall PCa, low- to intermediate-risk PCa, high- to very high-risk PCa and lethal PCa (defined in **Methods**), each compared to PCa-free controls (**Supplementary Tables 3**). After harmonizing metabolites (**Methods**), 595 associations were evaluated across the four PCa outcomes, representing 408 unique metabolites (101 amino acids, 8 carbohydrates, 13 cofactors and vitamins, 6 energy-related metabolites, 194 lipids, 20 nucleotides, 13 peptides and 53 xenobiotics) (**Supplementary** Figure 1). Of these associations, 149 (25%) were evaluated in ≥3 studies.

### Overall PCa

Nine studies (6,431 cases, 6,674 controls) reported on associations between 727 metabolite biomarkers and overall PCa risk (**Supplementary Table 3**), with 150 metabolites tested in ≥2 studies and therefore meta-analyzed (**Methods**). After applying an FDR (α=0.05) correction for multiple testing, three metabolites, amino acids serotonin and tiglylcarnitine (C5:1-DC) and the lipid sphinganine, were significantly associated with decreased overall PCa risk (**Figure 2, Supplementary** Figure 2**, Supplementary Table 4**). Nominal association evidence (unadjusted P<0.05) was observed for 25 metabolites (**Figures 2-3**, **Supplementary Table 4**). No metabolite pathway was significantly enriched among metabolites significantly or nominally associated with overall PCa risk (**Supplementary Table 5**, **Methods**).

**Figure 2:**
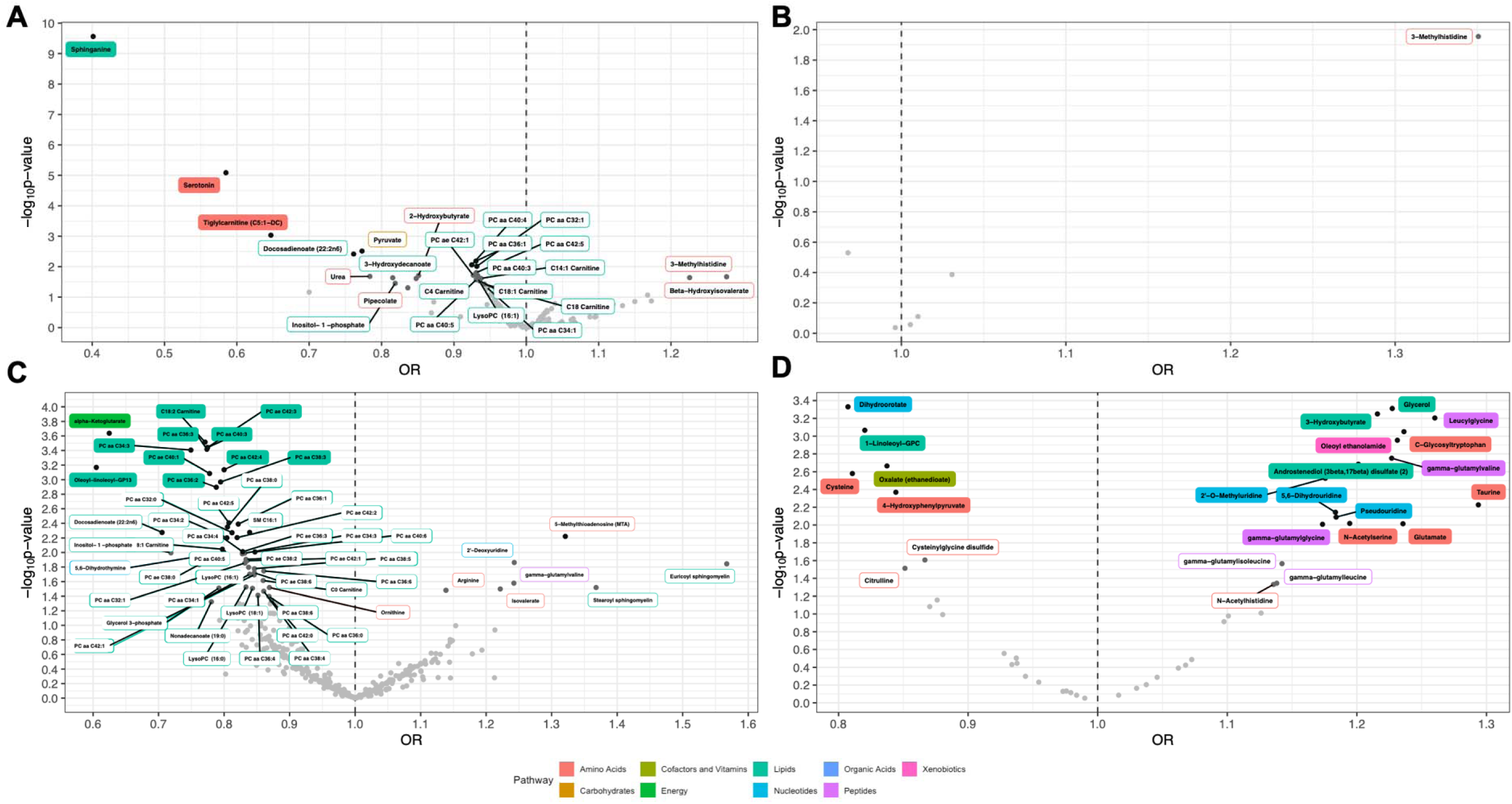
Meta-analysis association results between metabolites and PCa outcomes. Results are shown for A) overall PCa, B) low- to intermediate-risk PCa, C) high- to very high-risk PCa, and D) lethal PCa. Named metabolites were nominally associated (unadjusted P<0.05) with the respective PCa outcome. Shaded labels show FDR significant metabolites. Shading and label outline color represent metabolite class. Light grey point: P≥0.05. Dark grey point: P<0.05. Black point: P<0.01.

**Figure 3:**
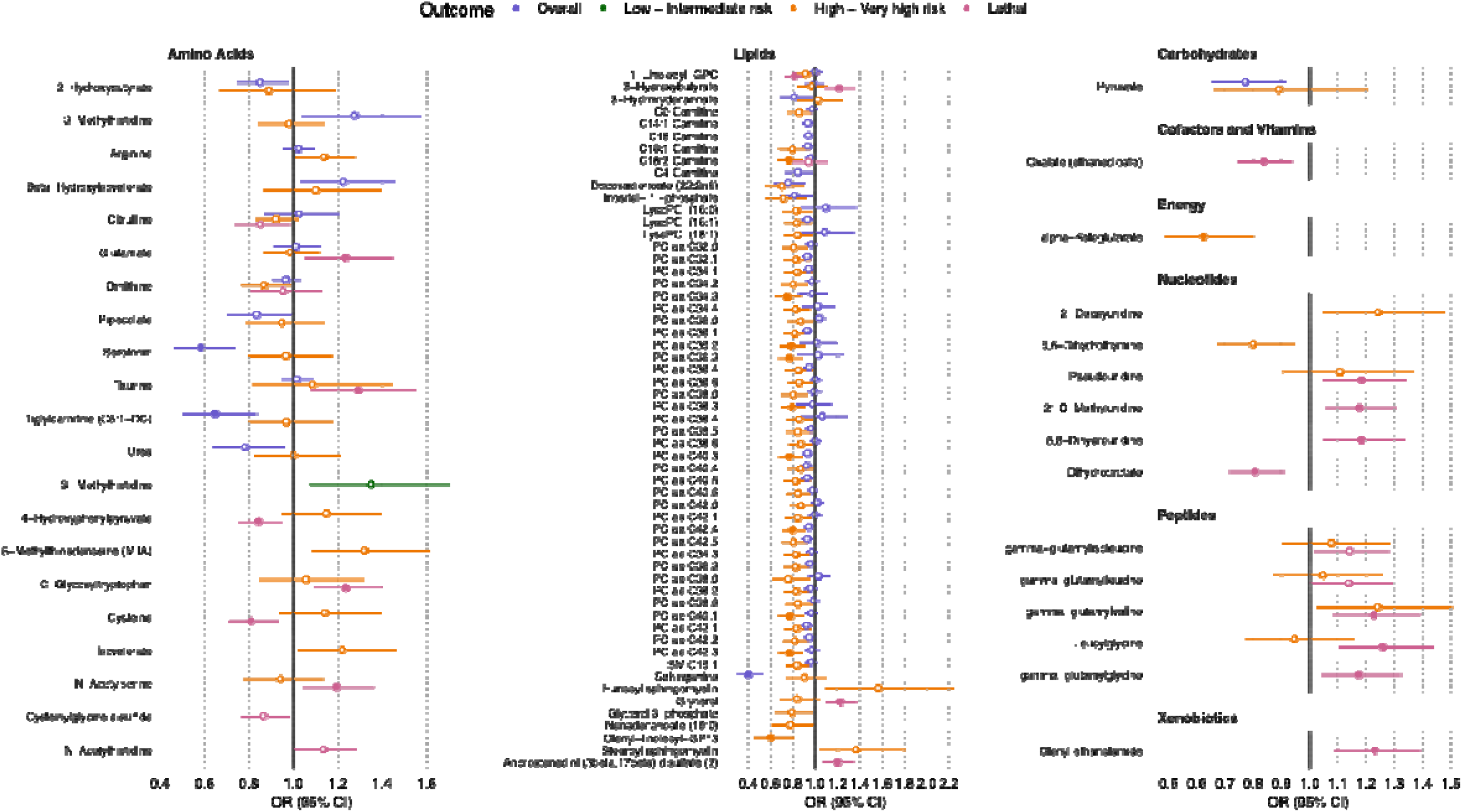
Association results for metabolites nominally associated with ≥ 1 PCa outcome. All tested outcomes are shown for each metabolite. Shaded circles indicate FDR significant associations.

### Low- to Intermediate-risk PCa

Four studies (3,454 cases, 4,505 controls) reported on associations between 186 metabolite biomarkers and risk of low- to intermediate-risk PCa (**Supplementary Table 3**), with six metabolites (all amino acids) tested in ≥2 studies and therefore meta-analyzed. No metabolite was significantly associated with risk of low- to intermediate-risk PCa (**Figure 2**, **Supplementary Table 6**). One amino acid, 3-methyl histidine, was nominally associated with increased risk of low- to intermediate-risk PCa (**Figures 2-3, Supplementary Table 6**). As all six metabolites were amino acids, enrichment analyses were not conducted.

### High- to Very High-risk PCa

Eight studies (1,470 cases, 6,679 controls) reported on associations between 1,001 metabolite biomarkers and risk of high- to very high-risk PCa (**Supplementary Table 3**), with 391 metabolites tested in ≥2 studies and therefore meta-analyzed. Eleven metabolites were significantly associated with decreased risk of high- to very high-risk PCa, including 10 lipids (9 phosphatidylcholines and 1 carnitine (C18:2 carnitine)) and 1 energy metabolite (alpha-ketoglutarate; **Figure 2**, **Supplementary** Figure 2, **Supplementary Table 7**). Fifty-five metabolites had nominal association evidence (**Figures 2-3**, **Supplementary Table 7**). Metabolites with significant and nominal association evidence were enriched for lipids (P_adj_=0.03 and P_adj_=5.71E-09, respectively; **Supplementary Table 5**).

### Lethal PCa

Four studies (1,059 cases, 4,999 controls) reported on associations between 806 metabolites and lethal PCa risk (**Supplementary Table 3**), with 48 metabolites tested in ≥2 studies and therefore meta-analyzed. Nineteen metabolites were significantly associated with lethal PCa risk, including positive associations with 14 metabolites (4 amino acids: C-glycosyltryptophan, taurine, glutamate and N-acetylserine; 3 lipids: 3-hydroxybutyrate, glycerol and androstenediol (3beta,17beta) disulfate (2); 3 nucleotides: 2’-O-methyluridine, 5,6-dihydrouridine and pseudouridine; 3 peptides: leucylglycine, gamma-glutamylvaline and gamma-glutamylglycine; and 1 xenobiotic: oleoyl ethanolamide) and inverse associations with 5 metabolites (amino acids cysteine and 4-hydroxyphenylpyruvate, cofactor/vitamin oxalate, phospholipid 1-linoleoyl-GPC and nucleotide dihydroorotate) (**Figure 2**, **Supplementary** Figure 2**, Supplementary Table 8**). Twenty-four metabolites had nominal association evidence (**Figures 2-3**, **Supplementary Table 8**). No pathway was significantly enriched among metabolites significantly or nominally associated with lethal PCa risk (**Supplementary Table 5**).

### Evidence of Effect Heterogeneity

Of the 595 associations tested, 280 (47.1%) had an I^2^=0, indicating the absence of between-study heterogeneity. The median I^2^ ranged from 0 for lethal PCa to 35.04 for low- to intermediate-risk PCa (**Figure 4**), with heterogeneity estimates largely comparable across metabolite pathways (**Supplementary Table 9**). In total, 94 (15.8%) associations had a Q-test P-value<0.05 and an I^2^≥40%, indicating between-study heterogeneity, with a lack of heterogeneity evidence predominantly observed for significantly (28/33, 84.85%) and nominally (81/105, 77.14%) associated metabolites (**Figure 4**).

**Figure 4:**
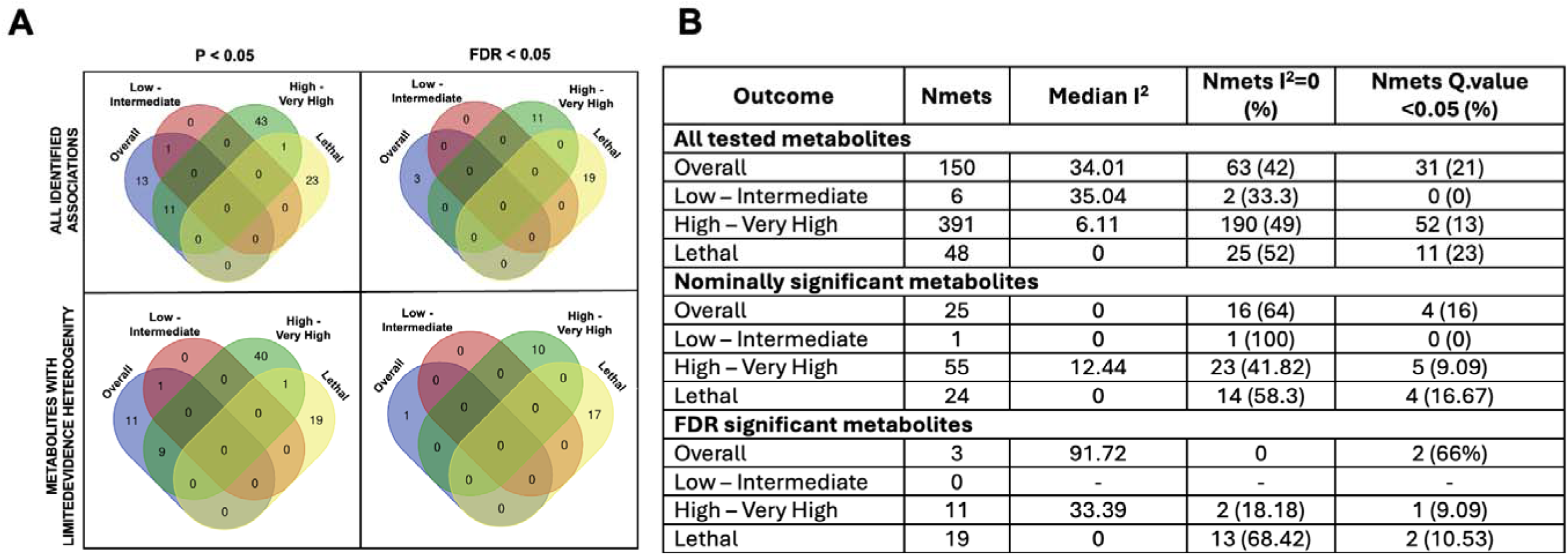
Summary of metabolite effect estimate heterogeneity by PCa outcome and significance threshold. **A**: Overlap of FDR significant and nominally significant metabolites before and after the removal of metabolites where N=2 some evidence of heterogeneity. **B:** Heterogeneity measures across all outcomes. Nmets: Number of metabolites.

Of the 150 metabolites evaluated with overall PCa, 28 were quantified with both MS and NMR, and four demonstrated between-platform heterogeneity (Q-test P-value<0.05). Of the 391 metabolites evaluated with high- to very high-risk PCa, 23 were quantified with NMR and MS, and one demonstrated between-platform heterogeneity. None of these five metabolites were associated with PCa outcomes **(Supplementary Table 10**). All metabolites evaluated with low- to intermediate-risk and lethal PCa were quantified with MS.

### Comparison of Evidence Between PCa Outcomes

No metabolite was significantly associated with >1 outcome; however, 13 metabolites were nominally associated with >1 outcome (**Figure 4**), all of which had consistent effect directions across outcomes. We found limited evidence of metabolite effects being correlated between outcomes, except for 11 metabolites nominally associated with both overall and high- to very high-risk PCa (R=0.91, P<1.1x10^-4^; **Figure 5**). Low- to intermediate-risk PCa was not considered in correlations, as only 6 metabolites were meta-analyzed for this outcome.

**Figure 5:**
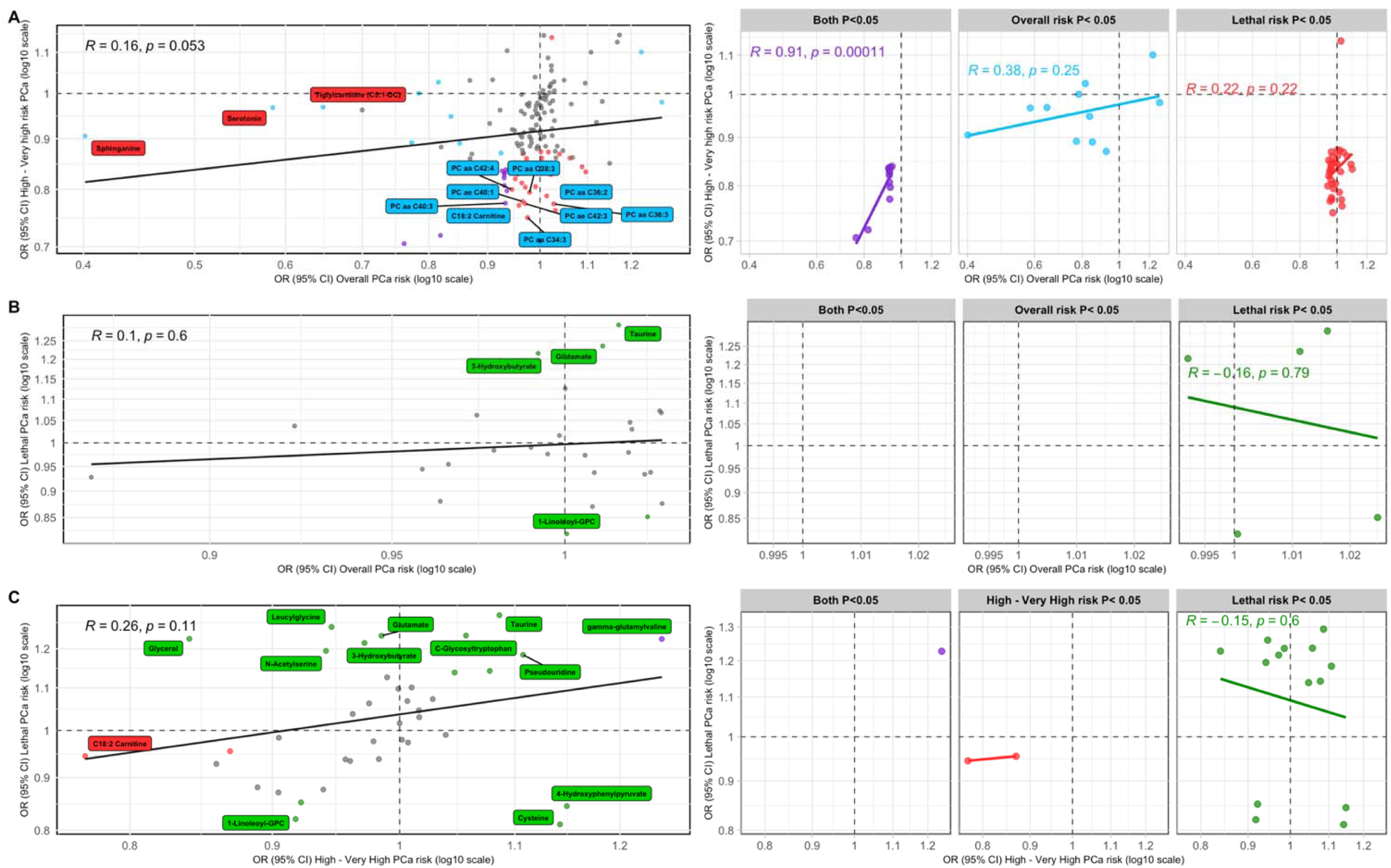
Pearson correlation of metabolite effect estimates between PCa outcomes. **A**: Overall vs high- to very high-risk PCa (N_mets_=143), **B**: Overall vs lethal PCa (N_mets_=27), **C**: High- to very high-risk vs lethal PCa (N_mets_=39). For each comparison, the three panels on the right indicate correlations between metabolites associated with an unadjusted P<0.05 in both traits or in one or the other trait. Purple points indicate results that were associated with both outcomes with an unadjusted P<0.05. Blue points indicate results that were associated with overall PCa with an unadjusted P<0.05. Red points indicate results that were associated with high- to very high-risk PCa with an unadjusted P<0.05. Green points indicate results that were associated with lethal PCa with an unadjusted P<0.05. Grey points indicate results that were associated with both outcomes with P≥ 0.05. Named metabolites were significantly associated (FDR<0.05) with the outcome of the given color. OR plotted on the natural log scale for clarity.

### Risk of Bias (ROB)

We assessed risk of bias (ROB) in our systematic review using a published tool that evaluates the quality of each included study based on six bias domains^24^ (**Methods**, **Supplementary Table 11**). Four of the 12 studies had low reporting quality (i.e., ROB score≤3), with bias most notably identified in the statistical analysis domain, primarily due to the selective reporting of significant findings in four studies (**Supplementary Table 12**). Bias in the exposure assessment domain, observed to some degree in six studies, was largely due to four studies not requiring fasting samples and one study reporting on m/z values rather than metabolite names, limiting the ability to interpret findings. Bias in the outcome domain occurred in six studies due to reporting the range/threshold of times between sample collection and PCa diagnosis rather than the median or mean. Additionally, seven studies were limited to participants that may not represent the general population, such as smokers or healthcare professionals. Participants predominantly represented European ancestries. To address these sources of potential bias, improve metabolite harmonization, and facilitate further metabolomic epidemiology meta-analyses, we have summarized key recommendations that should be considered in future metabolomic epidemiology studies (**Figure 7**).

### Associations Between Identified Metabolites and Other Cancers and Traits

We investigated whether any of the 33 total significant metabolites have been previously associated with other cancers and traits. Of these, 24 metabolites were identified in the Human Metabolite Database (HMDB)^25^, 17 of which had been previously associated with traits ranging from neurological conditions such as schizophrenia to diabetes and obesity, with 13 metabolites associated with ≥1 cancer type (colorectal, pancreatic, lung, breast and leukemia), although none were previously associated with PCa (**Supplementary Table 13**).

### Drug and Diet Targets

To explore whether the 33 PCa-associated metabolites were modifiable by diet, the FooDB and MetaboFood databases were searched. Twenty-one metabolites were identified in FoodDB, 8 of which were previously quantified in food, with the other 13 expected but yet to be quantified. Dairy products were the most common food identified, with 5 metabolites previously quantified in dairy products (tiglycarnitine, alpha-ketoglutarate, glutamate, pseudouridine, and taurine). One additional metabolite (oxalate, found in oranges) was identified in MetaboFood (**Supplementary Table 14**, **Figure 6**).

**Figure 6:**
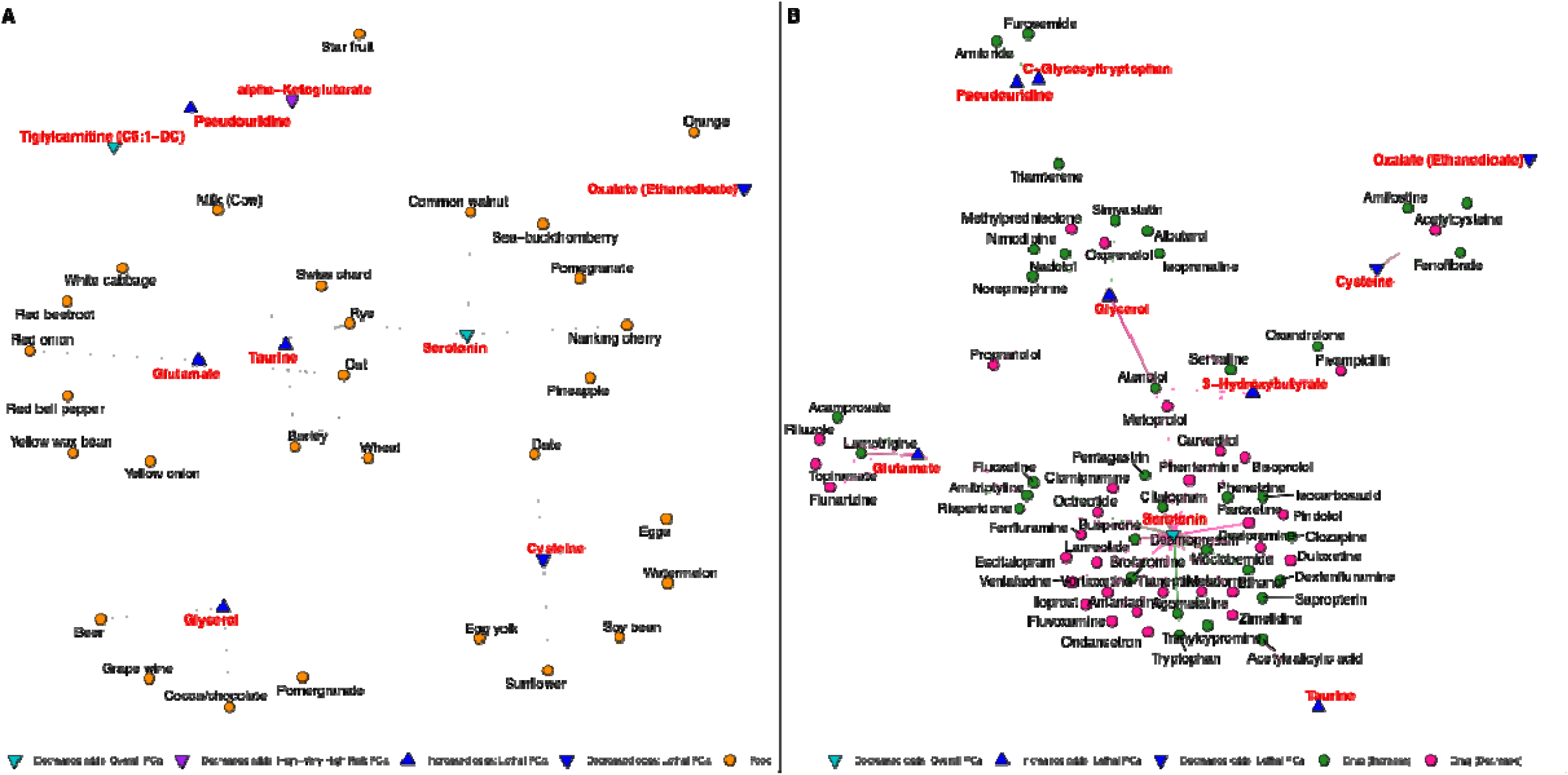
Identified dietary and drug modifiers of metabolites significantly associated with PCa outcomes. **A**: Foods that the PCa-associated metabolites have been previously quantified in according to FooDB and MetaboFood. Food names are harmonized for simplicity, with details provided in Supplementary Table 14. **B**: Drug modifiers of PCa-associated metabolites in the DrugBank pharmaco-metabolomics database; details are provided in Supplementary Table 15. PCa outcome associated with each metabolite is indicated by the color of the triangle, with upward triangles reflecting positive associations and downward triangles reflecting negative associations. In A, orange circles indicate foods, while in B, pink circles and lines indicate drugs that decrease metabolite levels and green circles and lines indicate drugs that increase metabolite levels.

**Figure 7:**
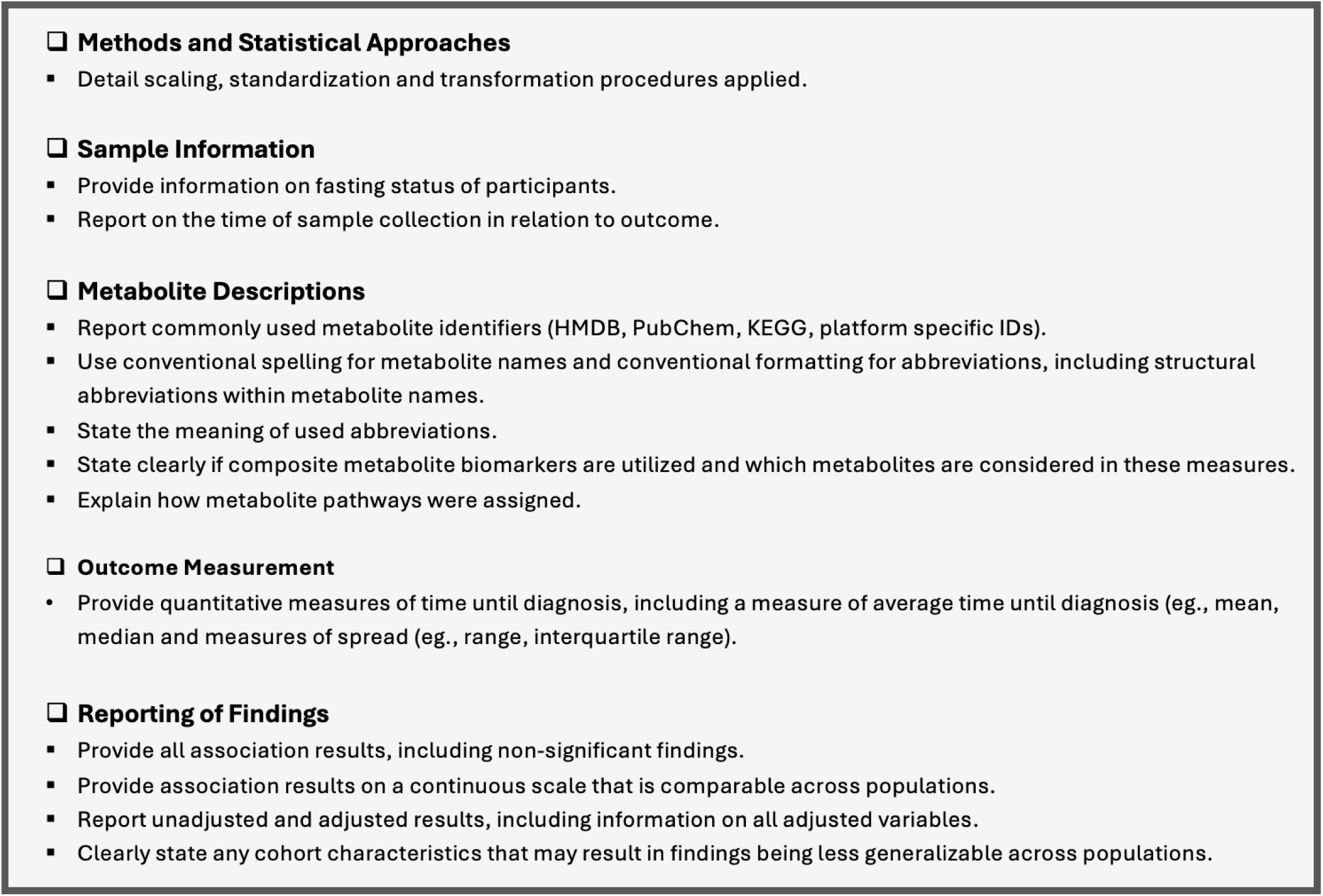
Recommended guidelines to improve reproducibility and facilitate meta-analyses of untargeted metabolomic epidemiology studies.

The DrugBank pharmaco-metabolomics database was searched to determine whether the PCa-associated metabolites were drug modifiable. Two metabolites associated with overall PCa risk were altered by drugs (sphinganine and serotonin), while 7 metabolites associated with lethal PCa risk were altered by drugs (taurine, C-glycosyltryptophan, pseudouridine, 3-hydroxybutyrate, glutamate, glycerol, and cysteine; **Supplementary Table 15, Figure 6**).

## Discussion

This large-scale systematic review and meta-analysis identified 33 circulating metabolites that were associated with PCa risk, with 3 associated with overall PCa, 11 associated with high- to very high-risk PCa, and 19 associated with lethal PCa risk. Metabolite risk profiles were unique to each PCa outcome, with no metabolite significantly associated with >1 outcome and metabolite effects not correlated between PCa outcomes. This suggests that distinct metabolite drivers may contribute to the development of more aggressive PCa, which could have notable implications for tailored risk stratification to discern risk of clinically significant PCa from indolent PCa.

Changes in tumor metabolism during cancer progression are well documented and result in metabolomic heterogeneity between cancer stages, providing an opportunity to identify distinct markers of advanced and lethal disease^26^. A metabolomic study of PCa biopsy tissue identified seven metabolites that significantly differed when comparing tumors with aggressive features (low Ki67, a cellular proliferation marker, and high PSA (>8ng/mL)) from less aggressive tumors (high Ki67 and low PSA), including the amino acid taurine and the ratio of glutamate/glutamine^27^. This corroborates our finding that taurine was associated with increased lethal PCa risk. Further, the long noncoding RNA taurine-upregulated 1 gene (lncRNA *TUG1*), a gene expression regulator that can be upregulated by taurine, was shown to be upregulated in prostate tumor tissue, with higher *TUG1* associated with more aggressive PCa, increased cell proliferation and poorer survival^28^, providing mechanistic insights into the role of taurine in lethal PCa.

We also found that glutamate was exclusively associated with lethal PCa risk, as suggested by previous systematic reviews^9,11^. Accordingly, a Mendelian randomization study found no evidence of association between glutamate and overall PCa risk^29^; however, higher serum glutamate has been reported in White and African American PCa patients with Gleason scores≥8 versus Gleason scores≤7 (measured post-diagnosis)^30^. Glutamate can be produced by the glutaminolysis pathway, converted into alpha-ketoglutarate, and then utilized in the tricarboxylic acid (TCA) cycle, increasing energy production in cancer cells and facilitating cellular proliferation^31^. Interestingly, we also identified a protective effect of alpha-ketoglutarate on high- to very high-risk PCa, which has been previously suggested^11^. Recent work has shown that sine oculis homeobox homolog 1 (SIX1), a newly identified PCa driver, contributes to migration and proliferation by acting as an upstream regulator of the enzyme glutamate-pyruvate transaminase 2 (GPT2), increasing cellular alpha-ketoglutarate production^32^. Future mechanistic work should further explore the role of glytaminolysis and the TCA cycle in PCa progression.

Amino acids are expected to contribute to PCa due to their essential role in maintaining redox balance, biosynthesis and homeostatic regulation^33^. Alongside taurine and glutamate, we found that amino acids C-glycosyltryptophan and N-acetylserine were associated with increased lethal PCa risk, while cysteine and 4-hydroxyphenolpyruvate (4-HPPA) were associated with decreased lethal PCa risk. C-glycosyltryptophan has been associated with increased all-cause and cardiovascular disease mortality^34^. Pre-diagnostic serum N-acetylserine has been associated with increased colorectal cancer risk^35^, and increased urinary N-acetylserine levels have been observed in endometrial carcinoma patients^36^. Cysteine, along with glutamate, is involved in the synthesis of glutathione (GSH), an antioxidant that helps mitigate oxidative stress from the accumulation of reactive oxygen species in cancer cells. *In vitro* and *in vivo* experiments found that intracellular L-cysteine depletion through the introduction of the engineered human enzyme Cyst(e)inase nearly depleted intracellular GSH leading to cancer cell death, reduced tumor growth in prostate and breast cancer xenografts, and increased DNA damage in PCa cells^37,38^, positioning Cyst(e)inase as a potential therapeutic agent. 4-HPPA is involved in tyrosine catabolism and is thought to reduce free radical levels, in turn reducing colon cancer risk^39^. As studies investigating the roles of these amino acids in PCa development are limited, future research may provide mechanistic insights into the potential for these to serve as preventive or therapeutic targets for clinically significant PCa.

Dysregulation of lipid metabolism is a characterizing feature of cancer^40^. Accordingly, we identified 1, 10 and 4 lipids that were significantly associated with risk of overall, high- to very high-risk and lethal PCa, respectively, with metabolites associated with high- to very high-risk PCa enriched for lipids. Nine of the 10 lipids associated with high- to very high-risk PCa were phosphatidylcholines (PCs), all of which were associated with reduced risk, with two also nominally associated with reduced risk of overall PCa. PCs are the most abundant membrane phospholipid and are precursors to other major membrane phospholipids (sphingomyelins and phosphatidylethanolamines) that can act as secondary messengers^41^. Due to this role in signaling and the generation of lipid mediators, PCs have been associated with both increased and decreased cancer risk^42^, with limited studies exploring how PCs link to PCa. We also found that glycerophospholipid 1-linoleoyl-GPC was protective against lethal PCa risk, a lipid that was previously associated with reduced kidney cancer risk^43^.

Three additional lipids, androstenediol (3beta,17beta) disulfate (2), 3-hydroxybutyrate (BHBA) and glycerol, were significantly positively associated with lethal PCa risk. Androstenediols are androgen steroid hormones that promote the growth of both normal and cancerous prostate cells through androgen receptor binding, and androgen deprivation therapy (ADT) is commonly utilized to treat metastatic hormone-sensitive PCa^44^. Unfortunately, most patients eventually develop ADT resistance, resulting in metastatic hormone-resistant PCa, which is responsible for most PCa deaths^45^. Hence, understanding the role of androgens in PCa development and progression is a major area of research. Androstenediol (3beta,17beta) disulfate (2) is a disulfated androgen, and although the role of steroid disulfates is not fully understood, the addition of two steroid groups is thought to reduce membrane transferability and receptor binding^46^. To understand if these could be targets for PCa prevention, future research is needed to understand the role of disulfated androgens in androgen signaling pathways that influence PCa progression through increased cellular proliferation, migration and decreased apoptosis^47^.

BHBA has been associated with increased and decreased cancer risk, likely due the ‘butyrate paradox’ in which some cancers preferentially utilize glucose as fuel and others preferentially oxidize BHBA^48^. BHBA acts as an inhibitor of cell proliferation via histone acetylation; therefore, in cancer types that preferentially oxidize BHBA, lower levels of BHBA are present and cell proliferation is not inhibited^48,49^. Indeed, in a cross-cancer meta-analysis of 1,900 metabolites, BHBA was one of the most upregulated blood metabolites across cancers^50^. In agreement with our findings, in a targeted multivariate serum metabolomics study, BHBA differentiated PCa cases with and without bone metastasis^51^. Furthermore, BHBA has been shown to increase proliferation and metastasis of colorectal cancer through the regulation of acetyl-CoA acetyltransferase (ACAT1)^52^, the expression of which has been postulated as a cancer therapeutic target and marker for PCa that may distinguish non-aggressive and aggressive disease^53,54^.

Glycerol is commonly used as food additive to introduce sweetness to foods. Although considered safe, our positive association with lethal PCa risk aligns with a study reporting that the rate of tumor growth of human PC3 hormone-resistant PCa cells was doubled in mice receiving glycerol compared to control mice receiving a saline solution, which was speculated to be a result of a reduction in oxidative damage^55^. Furthermore, glycerol is a by-product of glycerol-3-phosphate phosphatase (G3PP), an enzyme involved in glucose and lipid metabolism and the maintenance of redox balance. Elevated G3PP expression in PCa epithelial cells has been associated with increased risk of biochemical recurrence and bone metastasis, potentially due to G3PP alleviating metabolic stress in PCa cells^56^, demonstrating the potential prognostic utility of G3PP and the mechanistic relevance of glycerol to lethal PCa.

Four nucleotides were significantly associated with lethal PCa risk: pseudouridine, 2’-O-methyluridine, 5,6-dihydrouridine and dihydroorotate, with the latter three having positive associations and being related to pyrimidine metabolism. Limited studies have investigated the role of these nucleotides on PCa development; however, dihydroorotate is essential in the de-novo synthesis of pyrimidine and is the substrate for the enzyme dihydroorotate dehydrogenase (DHDOH), the inhibition of which is a potential target for cancer therapy due to the role of pyrimidines in RNA/DNA synthesis and cell proliferation^57^. Supporting the potential role of pyrimidine metabolism in lethal PCa, a previous systematic review^11^ and a prospective investigation that was ineligible for our meta-analysis (as it did not report risk estimates on a continuous scale) suggested a positive association between the pyrimidine nucleoside 2’-deoxyuridine and aggressive PCa risk^58^—consistent with our nominal evidence of association between 2’-deoxyuridine and risk of high- to very high-risk PCa. Future work is warranted to better understand the mechanistic role of pyrimidine nucleotides in lethal PCa and as potential biomarkers of clinically significant PCa.

Finally, three peptides were associated with increased lethal PCa risk: gamma-glutamylvaline, gamma-glutamylglycine and leucylglycine, with gamma-glutamylvaline also nominally associated with increased risk of high- to very high-risk PCa. Pre-diagnostic serum levels of these and several other gamma-glutamyl amino acids and dipeptides were associated with increased PCa-specific mortality in a survival case-only metabolomics investigation (that was ineligible for the current investigation)^59^. The enzyme responsible for the addition of a gamma-glutamyl group onto peptides, gamma-glutamyl transpeptidase (GGT), is expected to protect cancer cells against oxidative stress, leading to cancer cell growth and survival^60^. GGT has been shown to be upregulated in numerous cancers, including PCa, with greater levels observed in androgen-independent C4-2 and bone metastatic C4-2B cells compared to androgen-dependent LNCaP and partially androgen-independent C4 cells^61^. Further, expression of GGT was shown to increase tumor growth and chemoresistance in murine models^62^. Collectively, these findings demonstrate the importance of GGT in PCa progression and warrant additional research to understand how these peptides may act as biomarkers for lethal disease.

Our ROB assessment suggested that the publications included in this review were of good quality; however, key sources of bias were identified. One notable issue was the selective reporting of only significant results in three (25%) studies, which could contribute to publication bias and introduce false positive and false negative associations in our findings. In addition, inconsistent naming of metabolites and the reporting of associations based on spectroscopy measures (e.g., m/z values) could further bias our results due to the inability to accurately aggregate all reported associations. Inconsistent adjustment of covariates across studies was observed (**Supplementary Table 2**) and may have contributed to evidence of heterogeneity. The inconsistent reporting of findings stratified by time-to-diagnosis limited our ability to investigate temporal variability in metabolite-PCa associations, which has often been found to impact metabolite associations and will be important to investigate in follow-up studies of the metabolites reported here. Bias was also attributed to features of the primary cohort, including sample collection methods (e.g., collecting non-fasting samples, which could impact certain metabolite levels) and the generalizability of studies. Most notably, all identified studies were conducted in predominantly European descent individuals; therefore, our results may not be generalizable across populations, which is especially important given the stark health disparities of PCa, with Black and African American men having significantly higher incidence and mortality rates compared to White men^63,64^. This lack of diversity has also been noted in targeted metabolomic investigations of PCa^11^, further restricting the ability to validate metabolite findings across populations.

This is the first study to quantitively aggregate PCa metabolomic investigations to assess the relationship between untargeted circulating metabolites and clinically relevant PCa outcomes, combining the power of previous investigations into a large meta-analysis of ∼600 metabolites. With this approach, across the tested metabolites, we had 80% power to detect average relative risks of ≥1.23, 1.25, 1.51, and 1.29 for overall, low- to intermediate risk, high- to very high-risk, and lethal PCa, respectively, in fixed effects models (**Supplementary Table 16**). As such, this represents the most comprehensive understanding of metabolomic risk of PCa to date. Follow-up analyses of significant metabolites using drug and dietary databases also provides clinical and public health contexts into whether the identified metabolites could serve as dietary or drug targets.

However, this study has limitations. Due to the limited number of studies identified and overlap of metabolites between studies, we were unable to investigate potentially confounding factors (e.g., age or follow up time) via meta-regression or to formally assess publication bias. (e.g., funnel plot inspection or the harbord test). This also resulted in 75% of metabolites being tested in only two studies, restricting our ability to perform random effect analyses and more thoroughly assess heterogeneity. It will be important to further validate the PCa-associated metabolites only tested in two studies. Despite our extensive efforts to harmonize metabolites, naming discrepancies likely reduced the number of metabolites that could be successfully harmonized and meta-analyzed, particularly for poorly characterized metabolites and metabolites with multiple naming conventions (e.g., fatty acyls). Challenges in metabolite harmonization due to discrepancies in metabolite quantification, identification and annotations, along with the selective reporting of significant metabolite findings, have been similarly described in other metabolomic meta-analyses^50,65^ and will be important for the field to address to facilitate evidence synthesis from metabolomic studies and better understand the contribution of metabolites to disease risk. Towards this goal, we propose a set of guidelines for metabolomic epidemiology studies that could mitigate such challenges.

In conclusion, this study has identified a range of metabolites significantly associated with risk of clinically significant PCa. Metabolites associated with risk of high- to very high-risk and lethal PCa, such as glycerophospholipids, amino acids and pyrimidine nucleotides, may have clinical utility as biomarkers for early detection of PCa with poor prognosis. Future prospective investigations are warranted to assess the generalizability of these findings across diverse populations, particularly high-risk Black and African American populations. It also will be crucial to perform untargeted metabolomic studies across diverse populations to identify novel metabolite associations that may have been missed by previous investigations limited to predominantly European descent men. Further, conducting *in vivo* and/or *in vitro* experiments will be important next steps to determine the biological mechanisms underlying the metabolite associations reported here and better understand the potential for these metabolites to serve as biomarkers for PCa prevention, screening, and disease management.

## Methods

### Search Strategy

This systematic review is registered in the International Prospective Register of Systematic Reviews (PROSPERO; ID CRD42023462809). PubMed and Embase databases were systematically searched by one author (H.F.) for literature published through October 17^th^, 2024, using the following strategy: (’prostate cancer’ or ’prostate carcinoma’) AND (metabolite* or metabolomics or metabolomic) AND (serum or blood or plasma or circulating). Mesh indexing, human studies, English language, and original research filters were applied in each database, as applicable. Titles and abstracts were screened in duplicate by two authors (H.F. and O.P.A) using Abstrackr^66^, and disagreements were mediated by a third author (B.F.D.).

Inclusion criteria for the review were published original prospective human research studies written in English that reported on the association between PCa risk and ≥50 circulating metabolites (taken as an indication of an untargeted study) measured prior to prostate cancer diagnosis (i.e., cohort, nested case-control, or case-cohort). Randomized control trials (RCTs) were excluded due to their targeted nature. Given the accessibility of blood samples, which is ideal for biomarker development, and the commonality of blood-based metabolomic studies^4^, we limited studies to those conducted in blood samples. Studies that did not provide risk estimates for metabolites individually (rather they provided risk estimates for metabolite scores, conducted multivariate analyses, or evaluated prediction models) were also excluded along with studies that did not report metabolites on a continuous scale. Citation lists of included studies were searched until no additional studies were identified.

### Data Extraction and Synthesis

The following variables were extracted from included studies: first author, year, study name, study design, study population/country, case and control sample size, age at sample draw, years between sample draw and diagnosis, PCa definitions, sample type, fasting status, quantification technique, metabolomics platform, number of metabolites evaluated, main method of data analysis, model covariates, all metabolite identifiers, metabolite unit, effect estimates and standard errors. Odds ratios (ORs), risk ratios (RR), and hazard ratios (HRs) were interpreted as relative risks. Study authors were contacted to attempt to obtain missing results as needed.

When ≥1 publication was identified from the same cohort, the publication with the largest number of cases was selected unless it was stated that included individuals were independent from the previous publication (**Supplementary** Figure 3). When a study presented multiple effect estimates for a given metabolite, effect estimates adjusting for the most covariates were utilized. When effect estimates were only provided for specific strata (i.e., tumor stage, PTEN status, or age categories), the category most comparable to other studies included for the given outcome was selected to minimize heterogeneity.

When a study quantified a metabolite on both MS and NMR platforms, MS estimates were used in meta-analyses given the larger number of MS studies.

### PCa Outcome Definitions

Based on the case characteristics of included studies, four distinct PCa outcomes were assessed, based on NCCN definitions^23^: overall PCa, low- to intermediate-risk PCa, high- to very high risk PCa and lethal PCa, each compared to PCa-free controls (**Supplementary Table 4**). Overall PCa was based on national registries, cancer registries, and electronic health records. Due to limited sample sizes, “low- to intermediate-risk PCa” combined low-risk PCa (patients with grade group 1, cT1-cT2a staging and PSA<10ng/mL) and intermediate-risk PCa (patients with no high-risk features who had grade group 2-3, PSA of 10-20ng/mL and/or cT2b-cT2c staging). Likewise, “high- to very high-risk PCa” combined high-risk and very high-risk PCa (patients with cancer that had grown outside the prostate (cT3a or above), grade group of 4-5 and/or PSA>20ng/mL). Lethal PCa was defined as metastatic PCa or PCa-specific death. Studies defining PCa outcomes with other definitions were excluded to minimize heterogeneity.

### Metabolite Harmonization

Metabolites were harmonized based on COMP ID (when available for metabolites quantified by Metabolon), followed by HMDB IDs^25^ accessed online and via the R package hmdbQuery,^67^ PubChem IDs,^68^ and metabolite names. When metabolite names were used for harmonization, alternate metabolite names reported in HMDB and PubChem along with metabolite structures were also considered. Metabolite harmonization was verified using all available IDs and by manual check. Metabolites were mapped to biological pathways based on information provided in identified studies and HMDB.

### Statistical Analysis

Metabolites were meta-analyzed separately for each outcome using a restricted maximum likelihood (REML) approach for metabolites identified in ≥2 studies to reduce error resulting from meta-analyzing studies with small sample sizes^69,70^. Fixed effect meta-analysis results were presented when heterogeneity was limited (I^2^≤40%) or if there were two studies, with random effect meta-analysis results presented otherwise. Results were presented stratified by MS and NMR platforms to assess heterogeneity. Between-platform and between-study heterogeneity was assessed via the I^2^ statistic and a chi-squared test for subgroup differences (Q-test P-value <0.05). An FDR (α=0.05) correction was implemented to account for multiple testing separately for each cancer outcome. Nominal associations were defined as those with an unadjusted P<0.05.

To determine whether PCa-associated metabolites were enriched for any biological pathways, enrichment analyses were performed using a one-sided Fisher’s exact hypergeometric test with the R package bc3net^71^. Enrichment analyses were conducted on significant and nominally significant metabolites separately for each PCa outcome, with the reference panel including all metabolite meta-analyzed for a given outcome and pathway, comparing the number of observed to expected metabolites for a given pathway. Significance was defined using an FDR α=0.05 to account for multiple testing separately for each outcome.

We used Pearson correlation to determine how comparable effect estimates were between PCa outcomes for overlapping metabolites. This was conducted across all overlapping metabolites, metabolites with an unadjusted P<0.05 in both outcomes, and those with an unadjusted P<0.05 in one or the other outcome. Correlations were not assessed for outcomes with <15 metabolites due to insufficient data.

Results were reported following the Meta-analysis of Observational Studies in Epidemiology (MOOSE) and the Preferred Reporting Items for Systematic reviews and Meta-Analyses (PRISMA) guidelines^72,73^.

### Associations Between Identified Metabolites and Other Cancers and Traits

To evaluate whether any of the PCa-associated metabolites were previously associated with cancer or other traits, we searched the HMDB database on December 12^th^, 2024. Conditions listed under ‘associated disorders and diseases’ and the ‘health effects’ subcategory of physiological effects were extracted.

### Drug and Diet Metabolite Targets

To explore potential environmental modifiers of the identified significant metabolites, the FooDB and MetaboFood databases were searched to determine if these metabolites have previously been quantified in foodstuffs, and therefore, if any identified metabolites could potentially be dietary modifiable. Metabolites were searched using metabolite names (FooDB and MetaboFood), HMDB IDs (FooDB) and chemical structure (MetaboFood). To determine whether the PCa-associated metabolites could be drug modifiable, we searched the DrugBank pharmaco-metabolomics database^74^, utilizing all commonly used metabolite synonyms. Searches were conducted on July 29^th^, 2025.

### Risk of Bias (ROB) Assessment

Quality of the included studies was assessed with a published scoring tool^24^, previously utilized in other metabolomic epidemiology systemic reviews^70^, which evaluates six potential sources of bias: study participation, study attrition, exposure assessment, outcome assessment, evaluation of confounders, and the appropriateness of statistical analysis. Each domain was assigned a positive (‘yes’, scoring 1), neutral (‘somewhat’, scoring 0.5) or negative (‘no’, scoring 0) score to indicate whether the study avoided biases in each domain, based upon predefined criteria adopted for this review (**Supplementary Table 13**). An overall score was summated for each study, and a score ≤3 was considered low quality^70^. Question two on study attrition was not considered for nested case-control studies, as information on study attrition details were expected to be reported in the overall cohort study rather than in the nested case-control study. ROB was assessed by two reviewers (H.F and O.P.A).

## Additional Information

### Authors’ contributions

H.F. and O.P.A. performed the systematic review and B.F.D. mediated disagreements. H.F. and O.P.A. assessed the risk of bias criteria for each study. H.F. conducted all statistical analyses. J.M.G. assisted with public database searches and network figures. H.F. and B.F.D. drafted the manuscript. O.P.A. and J.M.G. provided critical review of the manuscript. H.F. and B.F.D contributed to study conception and design. B.F.D. supervised the investigation.

## Data Availability

All data produced in the present work are contained in the manuscript

