## Supplementary figures for "Pre-diagnostic circulating untargeted metabolomics and risk of overall and clinically significant prostate cancer: A systematic review meta-analysis"

**Supplementary Figure 1: Summary of included metabolites. A**: Overlap of investigated metabolites between investigated outcomes. **B**: Total number of metabolites tested for each metabolite pathway. **C**: Number of metabolites tested by pathway and outcome

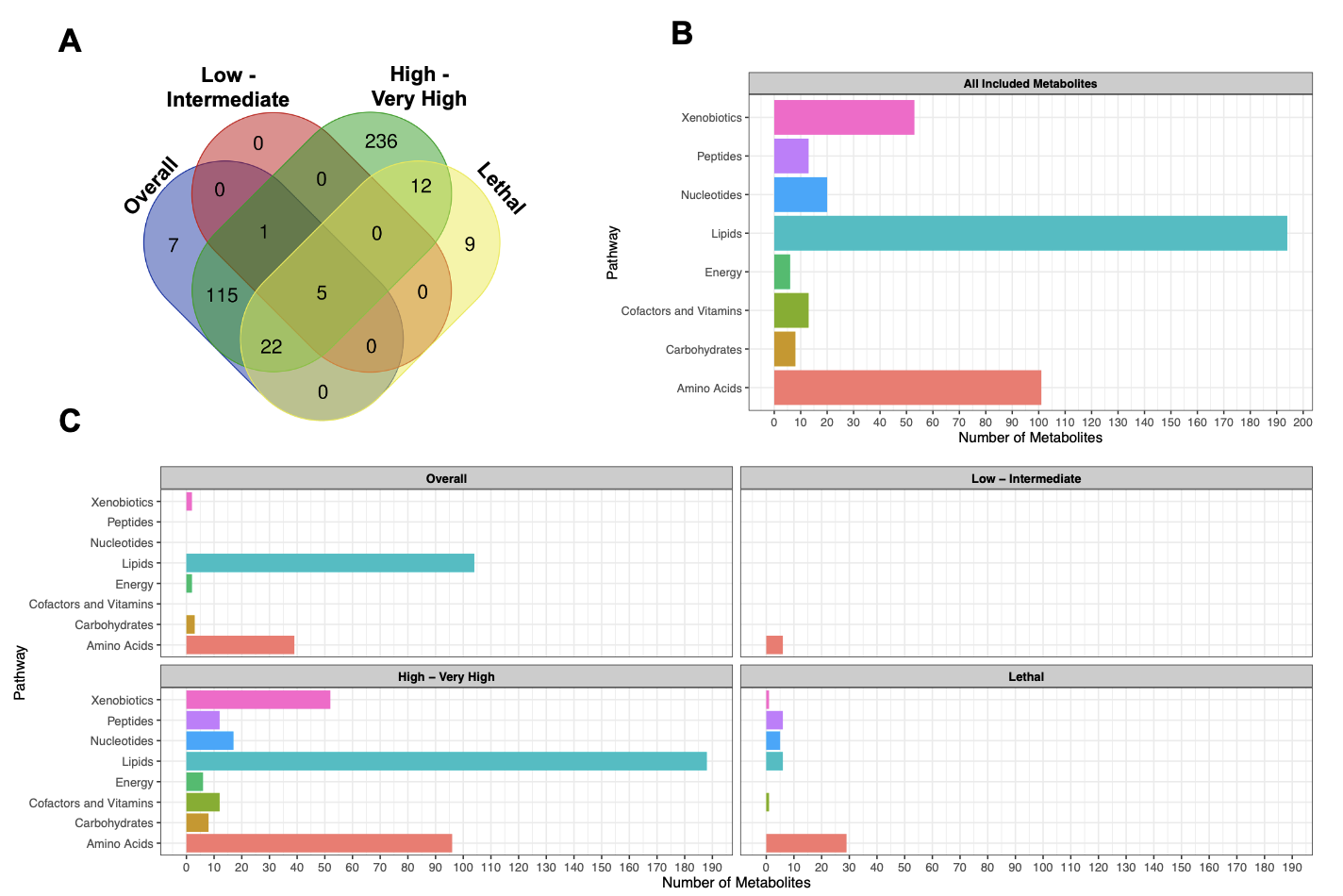

**Supplementary Figure 2: Meta-analysis forest plots for significant metabolite PCa- associations.** Yellow: Overall PCa. Orange: High-Very high risk PCa. Red: Lethal PCa

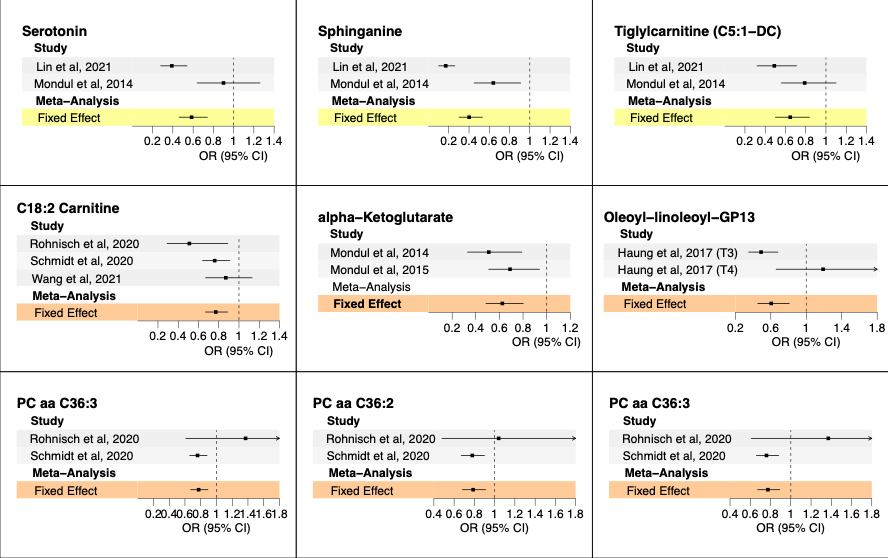

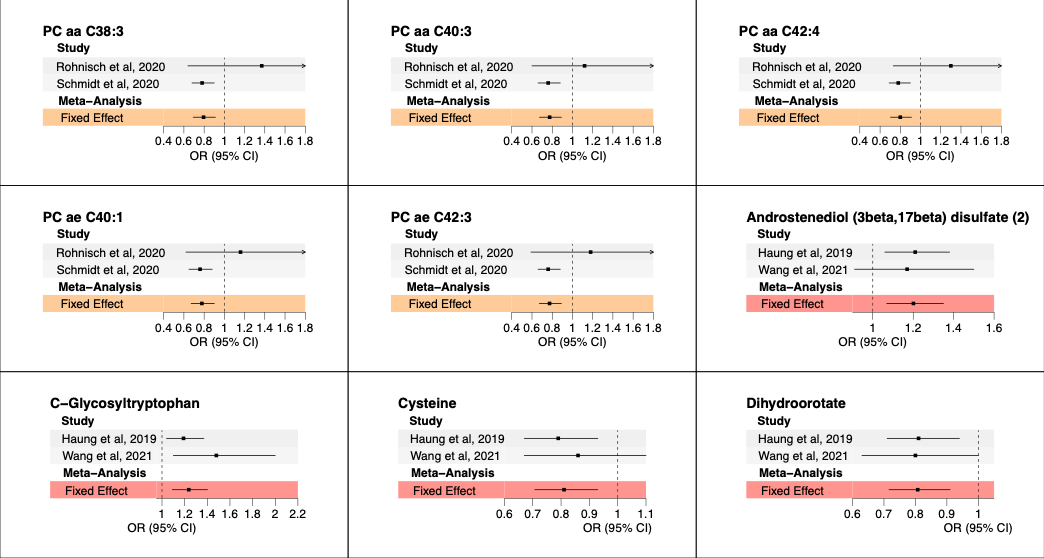

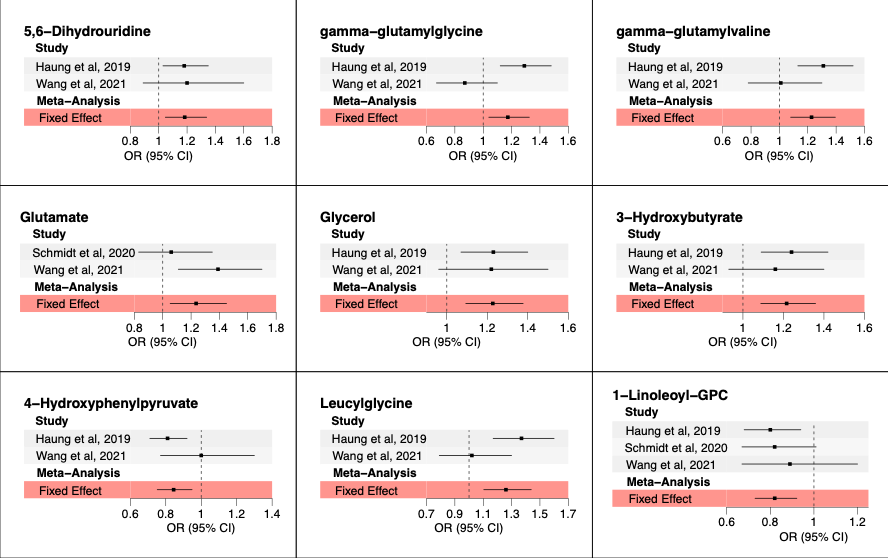

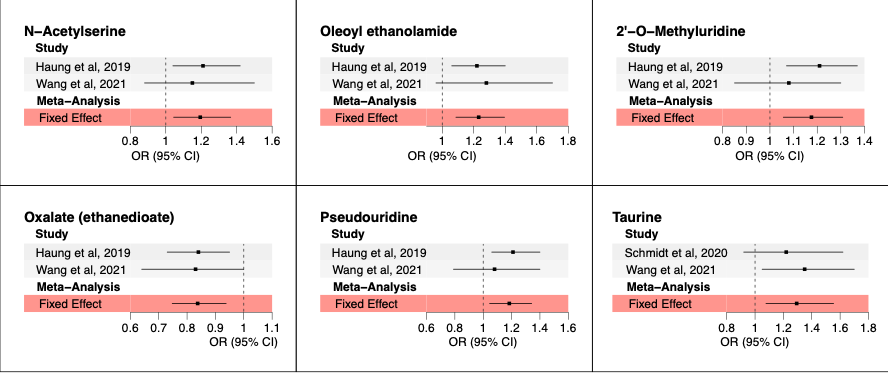

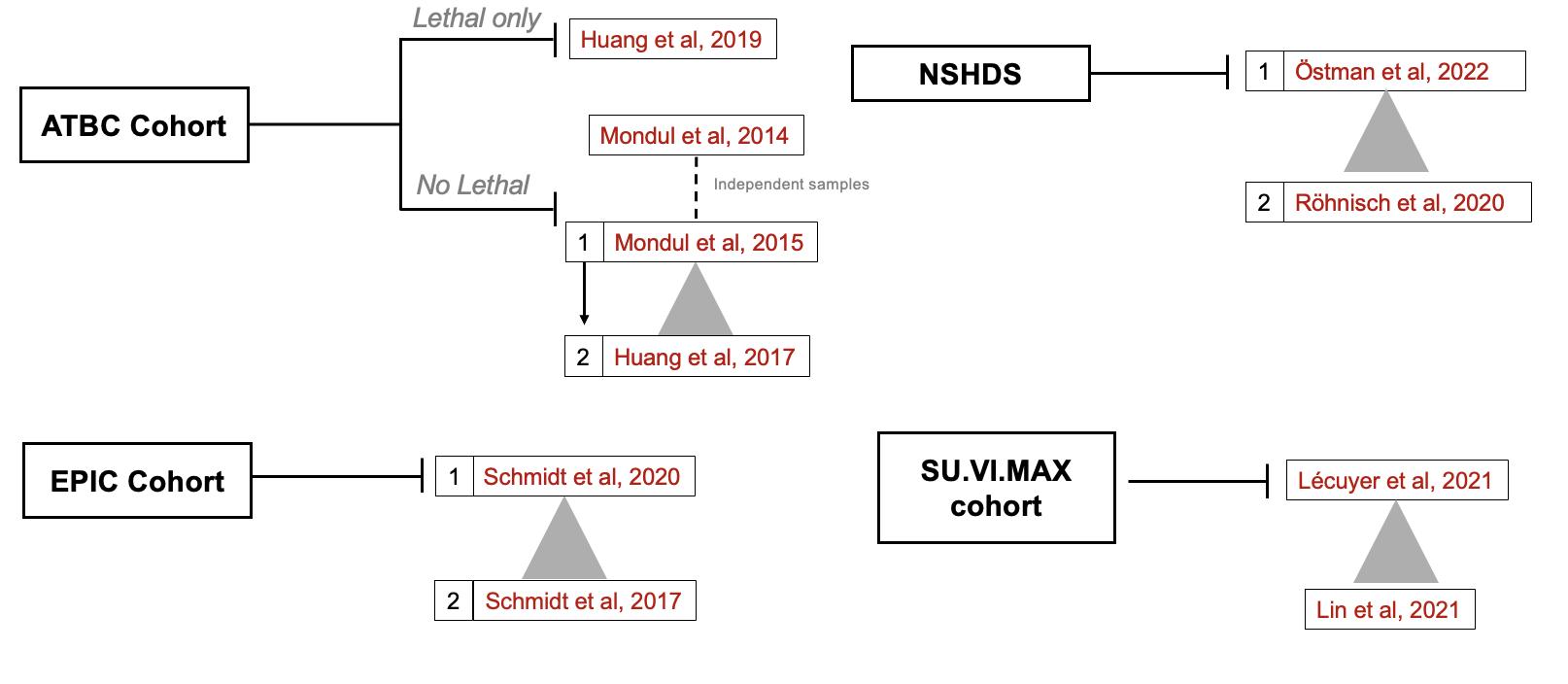
**Supplementary Figure 3: Study prioritization diagram when ≥1 study utilized potentially overlapping samples from the same cohort.** Prioritization was given to the study with the larger number of cases.
